# Differential DNA methylation in blood as potential mediator of the association between ambient PM_2.5_ and cerebrospinal fluid biomarkers of Alzheimer’s disease among a cognitively normal population-based cohort

**DOI:** 10.1101/2025.04.15.25325910

**Authors:** Tszshan Ma, Jiaqi Liu, Donghai Liang, Stefanie Ebelt, Kyle Steenland, Allan I. Levey, James J. Lah, Aliza P. Wingo, Thomas S. Wingo, Anke Hüls

**Affiliations:** Department of Epidemiology, Rollins School of Public Health, Emory University, Atlanta, GA, USA, 30322; Ganagarosa Department of Environmental Health, Rollins School of Public Health, Emory University, Atlanta, GA, USA, 30322; Department of Psychiatry, University of California, Davis, Sacramento, CA USA, 95816; Goizueta Alzheimer’s Disease Research Center, Emory University School of Medicine, Atlanta, GA USA, 30322; Department of Neurology, Emory University School of Medicine, Atlanta, GA USA, 30322; Division of Mental Health, Northern California VA, Sacramento, CA USA, 95816; Department of Neurology, University of California, Davis, Sacramento, CA USA, 95816; Alzheimer’s Disease Research Center, University of California, Davis, Sacramento, USA, 95816

**Keywords:** Alzheimer’s disease, DNA methylation, cerebrospinal fluid AD biomarkers, beta-amyloid plaques, fine particulate matter

## Abstract

**Introduction:** Fine particulate matter (PM_2.5_) is a known risk factor for Alzheimer’s disease (AD), with emerging evidence linking PM_2.5_ exposure to cerebrospinal fluid (CSF) biomarkers in pre-clinical stages. However, the role of DNA methylation (DNAm) as potential mediator in this relationship among cognitively normal individuals remains largely unexplored.

**Methods:** In 535 cognitively normal individuals, we assessed genome-wide blood DNAm, CSF Aβ_42_ concentrations, and residential PM_2.5_ exposure in the year preceding blood collection. Multi-stage comprehensive mediation analyses were conducted.

**Results:** Nine CpG sites mediated the PM_2.5_–Aβ42 association, with significant natural indirect effects (NIEs) for eight CpGs, mediating 14–43% of the effect. The joint NIE for all nine CpGs was -0.115 (95% CI: -0.215, -0.101) per 1 ug/m^3^ increase in PM_2.5_ exposure. Six CpGs are annotated to genes implicated in neuroinflammatory pathways.

**Discussion:** Our findings suggest that differential DNAm, particularly in neuroinflammation-related genes, mediates PM_2.5_ toxicity in AD’s pre-clinical stage.

## 1 Background

Alzheimer’s disease (AD), the most common form of dementia, is characterized by the progressive accumulation of beta-amyloid (Aβ) plaques and neurofibrillary tangles of hyperphosphorylated tau in the brain, leading to neuronal cell death and cognitive impairment[1]. With aging populations, AD has become a growing public health challenge[2]. The majority of AD cases occur in individuals over 65 and result from a combination of genetic predispositions, such as the apolipoprotein E (*APOE*) ε4 allele, and environmental exposures[3–5]. Increasing epidemiological studies have associated air pollution, particularly fine particulate matter (PM_2.5_), with increased risk of AD and related dementia[6, 7], and biomarkers of AD pathology, including Aβ and tau proteins[8–10]. However, the underlying biological mechanisms mediating these associations remain incompletely understood. Epigenetic regulation, particularly DNA methylation (DNAm), plays a critical role in neuronal function and cognitive health[11]. Growing research implicates DNAm in AD pathogenesis[12], with studies reporting DNAm alterations in key genes such as amyloid-beta precursor protein (*APP)* [13–15], presenilin 1 (*PSEN1)*[16], and *APOE*[13, 17] in postmortem brain samples. Epigenome-wide association studies (EWASs) have identified additional AD-associated loci, including *TMEM59*, which is involved in APP post-translational processing[18]; *DUSP22*, which may regulate tau phosphorylation[19]; *ANK1*, which plays a role in neuronal membrane compartmentalization[20, 21]; and the *HOXA* gene cluster, which contributes to embryonic development and neuroprotective function[22, 23]. Recently, meta-analyses of EWAS datasets validated previously identified genes and uncovered new differentially methylated positions related to inflammatory responses, DNA repair, and cell cycle regulation[24, 25].

Beyond its role in AD, DNAm is also influenced by environmental exposure such as PM_2.5_ exposure. Studies have reported that long-term PM_2.5_ exposure is linked to global DNA hypomethylation, potentially leading to genomic instability[26, 27]. PM_2.5_ exposure has also been associated with DNAm changes in inflammation-related genes [12], such as tumor necrosis factor (*TNF-a)* and interleukin-6 (*IL6)*, suggesting a role in promoting systemic inflammation[28, 29]. Similarly, PM_2.5_-induced DNAm changes in oxidative stress-related genes, such as glutathione S transferase (*GST*) and inducible nitric oxide synthase *(iNOS)*, may contribute to cellular damage and impaired detoxification pathways[28, 30]. EWASs have further identified PM_2.5_-related methylation alterations in loci involved in immune pathways, oxidative stress, and neuroinflammation, reinforcing the plausibility of epigenetic mediation in PM_2.5_-induced neurotoxicity[31–33]. However, formal mediation analyses assessing the causal role of DNAm in linking PM_2.5_ exposure to AD remain scarce.

We previously conducted the first human study linking traffic-related PM_2.5_, DNAm, and AD neuropathy using postmortem human brain samples from the Emory Goizueta AD Research Center (ADRC) brain bank[34]. Our findings suggested that differential DNAm in genes related to neuroinflammation, such as *AKT1S1* (Proline-rich AKT1 substrate 1), may mediate PM_2.5_’s effect on AD neuropathological markers in brain[34]. While DNAm in brain tissue offers the most direct evidence of AD-related epigenetic changes[12], blood-based analyses allow for earlier detection in cognitively normal individuals, enabling investigation of preclinical AD biomarkers and air pollution exposure in living populations[35].

Building on this foundation, the current study explored the relationship between ambient PM_2.5_ exposure, blood DNAm, and the AD cerebrospinal fluid (CSF) biomarker Aβ_42_ levels in cognitively normal participants from the Emory Healthy Brain Study (EHBS). We previously demonstrated that ambient PM_2.5_ is significantly associated with decreased CSF Aβ_42_ levels in this cohort[9]. To elucidate the underlying biological mechanisms—potentially involving epigenetic regulation and neuroinflammation—we examined whether differential DNAm in blood mediates the association between PM_2.5_ and CSF Aβ42 levels. To test this hypothesis, we applied a multi-stage analytical pipeline integrating traditional single-mediator analysis[36], high-dimensional mediation analyses, including DACT, HIMA1, HIMA2, HDMA, and MedFix [37–41], and causal mediation analysis for validation[42, 43]. Given the absence of a gold-standard approach for high-dimensional DNAm mediation analysis, this multi-method framework enhances the likelihood of identifying biologically meaningful CpG sites[44]. This study provides critical insights into the role of DNAm in mediating the effects of PM_2.5_ exposure on the pre-clinical stage of AD, shedding light on potential mechanisms relevant for intervention and prevention efforts.

## 2 Methods

### 2.1 Study Population

The study population for this analysis included participants in the Emory Healthy Brain Study (EHBS), a prospective research study focusing on the cognitive health of middle-aged and older adults. The EHBS is nested within the Emory Healthy Aging Study (EHAS) and includes participants 45−75 year of age who were free of cognitive impairment at enrollment from the Atlanta metropolitan region in the state of Georgia in the United States. More details on recruitment and eligibility can be found elsewhere[45]. The EHBS biennial study visits include neuropsychological tests, biospecimen collection (blood, CSF), cardiovascular measures, and brain imaging performed by trained clinical research staff[45]. Demographic characteristics of participants were collected with the online Healthy History Questionnaire (HHQ)[45]. The current cross-sectional analysis included data from the baseline visits conducted between 2016 and 2020. Of the 1,113 participants with CSF biomarker measurements, 536 also had genome-wide DNA methylation (DNAm) data available. The current analysis focuses on these 536 participants with both DNAm and CSF biomarker data. All participants provided informed consent, and the study received approval from the local ethics committee and the Emory University Institutional Review Board.

### 2.2 AD CSF Biomarker Concentrations

CSF biospecimens were collected by EHBS research staff via lumbar puncture at enrollment. CSF Aβ_42_ levels were quantified using the ElectroChemiLuminescense Immunoassay (ECLIA) Elecsys AD CSF portfolio on an automated Roche Diagnostics instrument (F. Hoffman-La Roche Ltd). The assay has measuring ranges of 200– 1,700 pg/mL for Aβ_42_. CSF Aβ_42_ level was normalized as z-score and kept as a continuous outcome in our main analyses.

### 2.3 Genome-wide DNA methylation measurement

Whole blood biospecimens were collected at enrollment and used for DNA extraction. DNAm measures were obtained with the MethylationEPIC BeadChips. The raw intensity files were transformed into a dataset that included beta values for each of the CpG sites, with values computed as the ratio of the methylated signal to the sum of the methylated and unmethylated signals, which ranged from 0 to 1 on a continuous scale. Pre-processing and statistics were done using R (v4.2.0). Following sample and probe filtering, 536 samples and 661,869 CpG sites remained for down-stream analyses. More details on the preprocessing and quality control have been published elsewhere[46]. The DNAm beta values were kept as a continuous mediator in our analyses. Whole blood cell type proportions (percentages of CD8+ T cells, CD4+ T cells, natural killer cells, B cells, monocytes, and neutrophils) was predicted using the most recent whole blood reference data set and the R package *minfi*[47, 48].

### 2.4 PM_2.5_ Exposure Assessment

High-resolution ambient PM_2.5_ exposure data was obtained from the publicly available Socioeconomic Data and Application Center (SEDAC) air quality dataset for health-related applications[49]. The data consists of yearly average ambient PM_2.5_ concentrations (in μg/m^3^) estimated at a 1 km × 1 km resolution for the entire contiguous United States (2000–2016). The PM_2.5_ predictions were derived using established spatiotemporal ensemble models that integrated three different machine learning algorithms (neural network, random forest, and gradient boosting machine) and included over one hundred predictors from satellite measurements, land-use terms, meteorological variables, and chemical transport model predictions[50]. The quality of the estimates was assessed using 10-fold cross-validation against monitoring measurement values from the Environmental Protection Agency (EPA) Air Quality System across the United States. The resulting R^2^ values for national annual predictions were 0.89[50]. As our study participants all lived in the state of Georgia, we restricted the PM_2.5_ estimates to Georgia. Participants’ geocoded residential addresses at their baseline visit were mapped using QGIS mapping software and we spatially matched geocoded residential addresses to the closest centroid of 1-km^2^ PM_2.5_ grids to assign annual exposures at the year for specimen collection[9].

### 2.5 Covariates

Individual-level demographic characteristics (self-reported gender, age, race/ethnicity, educational attainment, and body mass index (BMI)) were obtained from the online Health History Questionnaire[45]. Self-reported gender was a binary variable: female, male. Age as a continuous variable denotes the age at the year of visit for biospecimens collection. Because of the presence of multi-ancestral groups and small categories in self-reported race/ethnicity, we used a three-level race variable: White, Black/African American, and Other, as well as a binary ethnicity variable indicating Hispanic origin. Similarly, educational attainment was included as a three-level variable: less than a college degree, college degree, master’s degree or higher. Height and weight measurements were used to calculate BMI (weight in kilograms divided by height in square meters) as a continuous variable. We also included two area-level characteristics, Area Deprivation Index (ADI) and principal components of neighborhood deprivation, to denote participant’s neighborhood socioeconomic status (SES). The ADI is provided in national percentile rankings at the census block group level from 1 to 100, where 100 represents the most deprived neighborhood, and was calculated using census block group-level indicators and factor analysis to cluster indicators based on their ability to explain the variance between block groups[51]. The three principal components of neighborhood deprivation were calculated based on estimates from American Community Survey (ACS) census tract-level data, including 15 indicators of 6 socioeconomic domains (poverty/income, racial composition, education, employment, occupation, and housing properties) (see Li et al. 2022[52] for details). We assigned each participant’s ADI, and three principal components of neighborhood deprivation variables based on their residential address[9]. In addition to individual-level and area-level demographic covariates, we also include whole blood cell-type proportions as covariates due to their substantial impact on DNA methylation. The confounding structure was determined according to literature review and our previous studies[9], and is illustrated by directed acyclic graphs (DAGs) in **Figure S1**.

### 2.6 Statistical Analysis

#### Total Effect Analysis

Before conducting the mediation analysis, we assessed the total effect of PM_2.5_ exposure on CSF Aβ_42_ concentrations without considering DNAm as a mediator. Previously, we found higher PM_2.5_ exposure was associated with decreased CSF Aβ_42_ levels among 1,113 participants in the EHBS cohort, suggesting an accumulation of amyloid plaques in the brain and an increased risk of developing AD [9]. We repeated this total effect analysis among the subset of 536 participants with DNAm data with the following linear regression model:

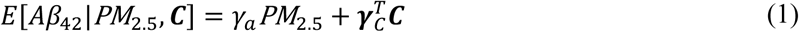

Here, *PM*_2.5_ is the continuous exposure variable, *Aβ*_42_ is the continuous outcome as the CSF Aβ_42_ z-score, and ***C*** denotes the adjustment set of covariates, including sex, age, race/ethnicity, educational attainment, BMI, ADI, three principal components of neighborhood deprivation, as shown in the DAG (**Figure S1A**).

#### Mediation Effect Analysis

Next, we aimed to identify blood DNAm patterns that potentially mediate the association between PM_2.5_ exposure and decreased CSF Aβ_42_ levels. Given the lack of consensus in the literature regarding best practices for conducting mediation analysis with high-dimensional mediators[53], we developed a comprehensive analysis pipeline similar to our previous work [34, 54, 55] (**Figure 1**; see **Figure S2** for additional details on each method). We began by conducting a traditional single-mediator mediation analysis using the product method to assess each CpG site individually and screen for CpG sites potentially associated with both PM_2.5_ exposure and decreased CSF Aβ_42_ levels[36]. Next, we applied five high-dimensional mediation analysis methods (DACT, HIMA1, HIMA2, HDMA, and MedFix) to identify noteworthy mediators among the CpG sites filtered in the first step [37–41]. These high-dimensional methods encompassed a diverse array of statistical approaches, increasing the likelihood of identifying relevant mediators[44]. Finally, we validated the noteworthy CpG sites identified in the second step by performing causal mediation analysis to estimate the natural indirect effects of each CpG site and the joint indirect effect of all noteworthy CpG sites [42, 43]. All analyses were completed in R (v4.4.1).

**Figure 1.**
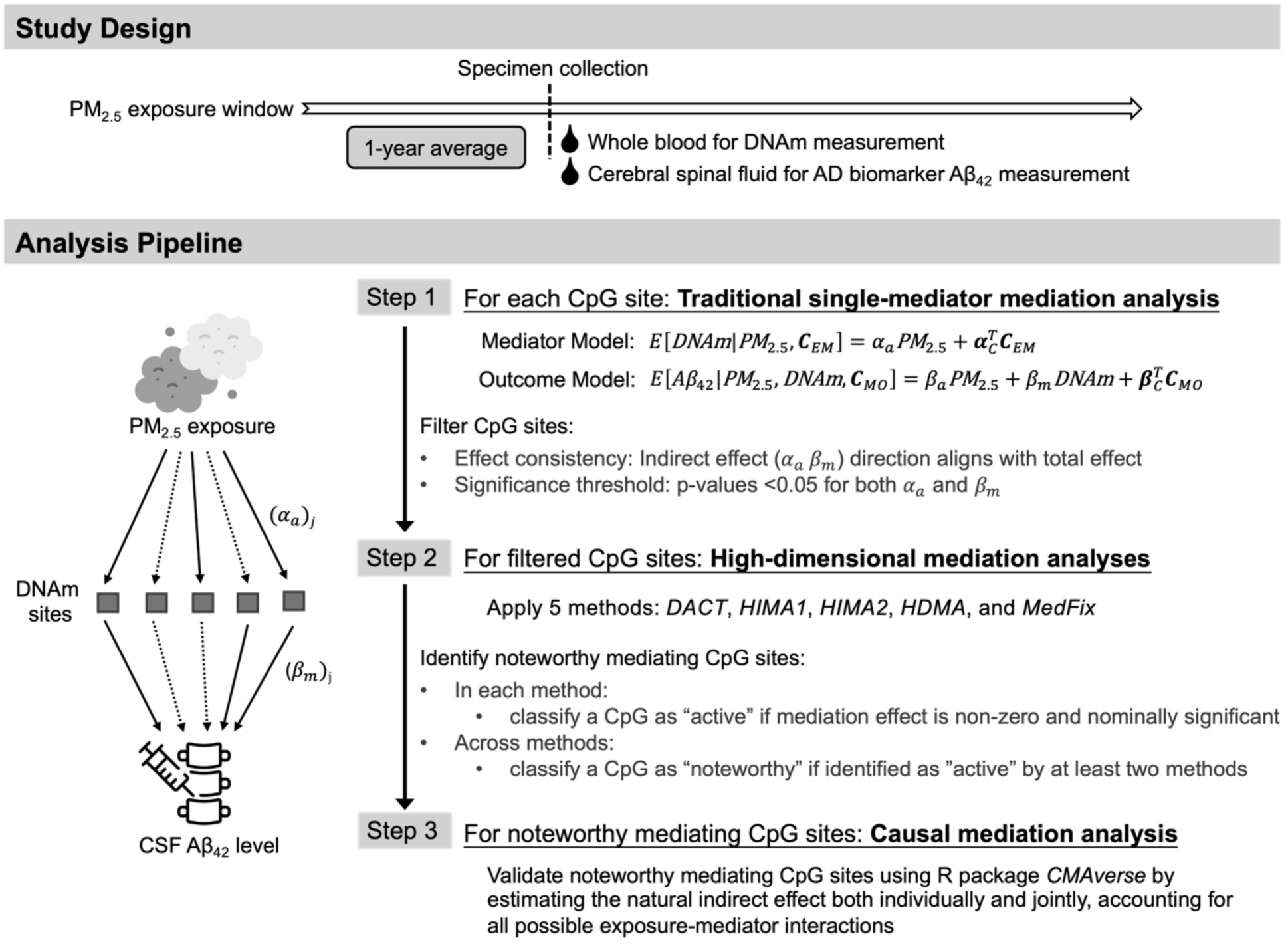
Graphical overview of the study design and the analytical pipeline used to identify and evaluate potential mediating CpG sites in the association between PM_2.5_ (exposure) and CSF AD biomarker Aβ_42_ concentrations (outcome). The pipeline incorporates single-mediator analysis, high-dimensional mediation analysis, and causal mediation analysis. Note: PM_2.5_, fine particulate matter; CSF, cerebral spinal fluid; AD, Alzheimer’s disease; Aβ_42_, beta-amyloid 42; ***C***_EM_ and ***C***_MO_ denote the covariate matrices for the mediator model and outcome models, respectively.

#### Step 1: Traditional single-mediator mediation analysis

We first assessed the CpG site “one-at-a-time” using the standard regression-based approach proposed by Baron and Kenny (1986)[36]. The following robust linear regression models were fitted for each CpG site:

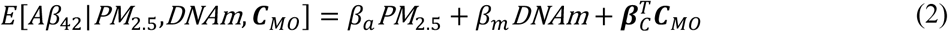

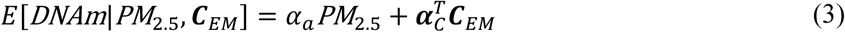

to estimate the mediating role of *DNAm* in the causal pathway between the exposure *PM*_4.5_ and outcome *Aβ*_4 2_[56]. Model (2) represents the outcome model, while Model (3) represents the mediator model. Here, *PM*_4.5_ is the continuous exposure variable, *Aβ*_42_is the continuous outcome as the CSF Aβ_42_ z-score, and *DNAm* the continuous mediator as the DNAm beta value. Covariates in the adjustment sets ***C**_MO_* (for the outcome model) and ***C**_EM_* (for the mediator model) included sex, age, race/ethnicity, educational attainment, BMI, ADI, three principal components of neighborhood deprivation, and proportion of blood cells, as shown in the DAGs for the mediator and outcome models (**Figure S1B** and **S1C**). To identify the natural indirect effect (NIE) mediated through DNAm, the following assumptions need to be met[57, 58]: (i) no unmeasured exposure-outcome confounding conditional on ***C***; (ii) no unmeasured mediator-outcome confounding conditional on (***C***, *PM*_4.5_); (iii) no unmeasured exposure-mediator confounding conditional on ***C*** ; (iv) no effect of exposure that confounds the mediator-outcome relationship; and (v) no exposure-mediator interaction on the outcome. Under these standard assumptions, the NIE is *⍺_a_β_m_*, the direct effect of exposure on outcome is *β_a_*, and the total effect is *β_a_* + *⍺_a_β_m_*.

We fitted the above robust linear regression models for each CpG site using R packages *MASS*, *lmtest*, and *sandwich*. Only CpG sites with p-values <0.05 for both the exposure-mediator association (*⍺_a_*) and the mediator-outcome association (*β_m_*) were included in downstream analyses. Furthermore, we retained only CpG sites with negative indirect effects (*⍺_a_β_m_*< 0) to align with previously observed direction of the total effect[9]. This pre-screen step strategically reduced dimensionality of the data, facilitating the feasibility of subsequent high-dimensional mediation analysis[54, 55].

#### Step 2: High-dimensional mediation analysis

We applied five high-dimension mediation analysis methods—DACT, HIMA1, HIMA2, HDMA, and MedFix—to identify noteworthy mediators among the filtered CpG sites. Since these methods are relatively novel, there is currently no established gold standard for high-dimensional mediation analysis[53]. DACT (*divide-aggregate composite null test*) by Liu et al. (2022) utilized the Efron empirical null framework to calculate a weighted sum of p-values obtained from the outcome and mediator models (Model 2 and 3) to test for the significance of mediators[37]. The remaining four methods (HIMA1, HIMA2, HDMA, and MedFix) extended Model (1) and (2) by fitting multiple mediators simultaneously using penalized regression for variable selection and effect estimation[44]. HIMA1 (*high-dimensional mediation analysis*) by Zhang et al. (2016)[38] employed a dimension reduction technique based on sure independence screening (SIS) using p-values from the single-mediator outcome model[59]. It then applied the minimax concave penalty (MCP)[60] for mediation effect estimation and joint significance testing, assuming a uniform null distribution for p-values[61, 62]. HDMA (*high-dimensional mediation analysis*) by Gao et al. (2019)[40] developed along the lines of HIMA1 but replaced the MCP with the de-biased LASSO penalty[63] for mediation effects estimation. HIMA2, developed by Perera et al. (2022), was an extended version of HIMA1[39]. Like HDMA, HIMA2 also utilized the de-biased LASSO penalty for effect estimation but improved the SIS approach by ranking CpG sites based on the absolute magnitude of estimate indirect effect (αβ). Additionally, it implemented a p-value correction procedure that allowed a mixture of null distributions[64]. MedFix (*mediation analysis via fixed effect model*) by Zhang (2022) extended HIMA for settings involving multiple exposures and multiple mediators[41]. It employed adaptive LASSO for effect selection in both the outcome and mediator models[65]. Although MedFix was designed for multi-exposure scenarios, in our single-exposure setting, it functioned similarly to HDMA but used adaptive LASSO instead of debiased LASSO. We conducted DACT analysis using the R package *DACT*, while HIMA1, HDMA, and MedFix analyses were performed using the R package *hdmed*. HIMA2 analysis was carried out using the R package *HIMA*.

In each separate high-dimensional mediation analysis, we classified a CpG site as “active” if its estimated mediation contribution was non-zero and its raw p-value was <0.05. Following the recommendation in Clark-Boucher et al. (2023), we considered “active” CpG sites identified by at least two methods as noteworthy mediators[44].

#### Step 3: Causal mediation analysis

Noteworthy CpG sites were validated using causal mediation analysis performed with the R package *CMAverse*, which provided estimates of NIE, natural direct effect (NDE), total effect (TE), and proportion mediated (PM)[42, 43, 58]. We assessed mediators under two scenarios: (1) individually, allowing for exposure-mediator (E-M) interactions during effect estimation, and (2) jointly, considering all noteworthy CpG sites together while also allowing for E-M interactions. This multi-mediator approach relaxed the assumption that there were no mediator-outcome confounders affected by the exposure, allowing for more realistic modeling where one mediator might influence another within the causal pathway[66]. This approach enhanced the biological plausibility of the estimates by reflecting the interconnected nature of CpG sites within biological pathways. The 95% confidence intervals for effect estimates were constructed using bootstrapping, and two-sided p-values were reported.

### 2.7 Sensitivity analyses

Given that CSF AD biomarker concentrations may differ by race[67], we repeated the causal mediation analysis for the identified noteworthy CpG sites, restricting the study population to White participants, who comprised 85.4% of our cohort (see **Table S1** for descriptive characteristics of the White EHBS participants).

### 2.8 Secondary analyses

To further support our findings, we conducted a blood-brain concordance analysis for CpG sites identified as noteworthy mediators in the high-dimensional mediation analyses (step two). We assessed the correlation between DNAm in blood and brain tissue using the Blood-Brain Epigenetic Concordance (BECon) tool[68] and the Gene Expression Omnibus Database [Accession code GSE111165]. Additionally, we performed a gene ontology (GO) functional enrichment analysis using the R package *missMethyl*[69], based on the broader set of CpG sites filtered from the standard single-mediator mediation analysis in step one, which were associated with PM_2.5_ exposure and decreased CSF Aβ_42_ levels. All CpG sites were annotated using the annotation data for the “IlluminaHumanMethylationEPIC” array[70]. For further functional insight into individual CpGs sites and their corresponding annotated genes, we searched previous studies using the EWAS catalog[71] and conducted literature searches on PubMed.

## 3 Results

### 3.1 Study Population Characteristics

The analysis included 536 EHBS participants with available CSF AD biomarker data, whole blood DNAm and other relevant covariates (**Table 1**). The average age of the study population was 61.8 years (standard deviation [SD] = 7.0), with 70.3% identifying as female. The majority of participants (85.4%) identified as White, 9.5% as Black/African American, and 2.8% as Hispanic. The average BMI was 25.5 kg/m^2^ (SD = 3.7). Most participants, 449 (83.8%), completed college or higher education, and lived in less deprived neighborhoods compared to the national average (ADI: mean = 29.7, SD = 20.4). As shown by the 1-year average residential PM_2.5_ exposure map (**Figure 2**), participants residing in the southern areas of the Atlanta city experienced higher PM_2.5_ levels compared to those living in northern communities. The mean of 1-year ambient PM_2.5_ exposure was 9.53 μg/m^3^ (interquartile range [IQR] = 0.79), slightly above the latest annual National Ambient Air Quality Standards (NAAQS) PM_2.5_ threshold of 9 μg/m^3^[72]. CSF Aβ_42_ concentrations displayed considerable variability within the study population (median = 1230.5 pg/mL, IQR = 640.8) (**Table 1**). While CSF Aβ_42_ concentrations followed an approximately normal distribution, there was a notable frequency peak at the highest levels, indicating that most participants had Aβ_42_ concentrations within the normal range (**Figure S3**).

**Figure 2.**
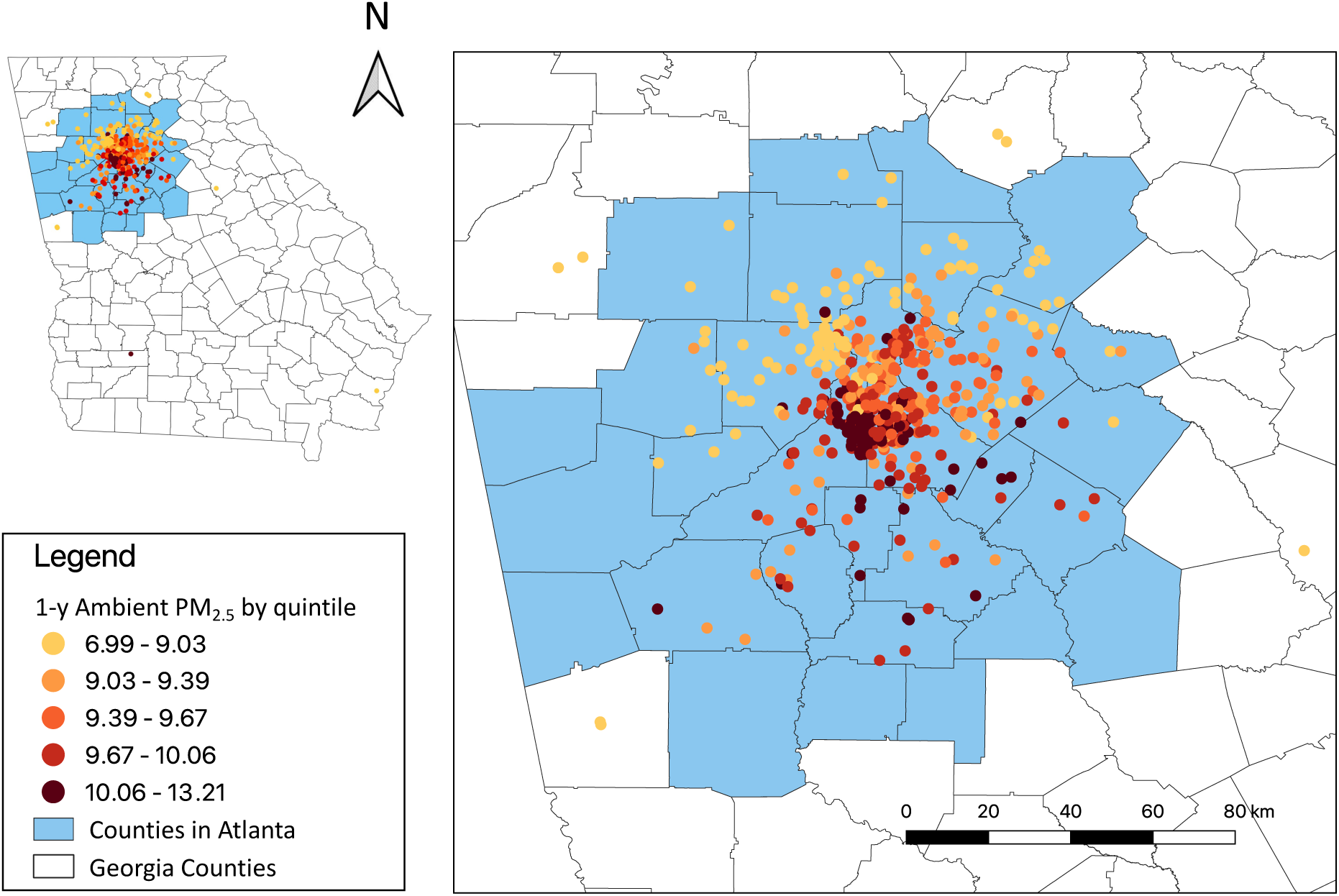
Geographic distribution of the study population and their residential PM_2.5_ exposure concentrations in the year prior to specimen collection. Each dot represents a participant (n = 536) and their 1-year averaged ambient residential PM_2.5_ exposure (in ug/m^3^), categorized by quintiles.

**Table 1.**
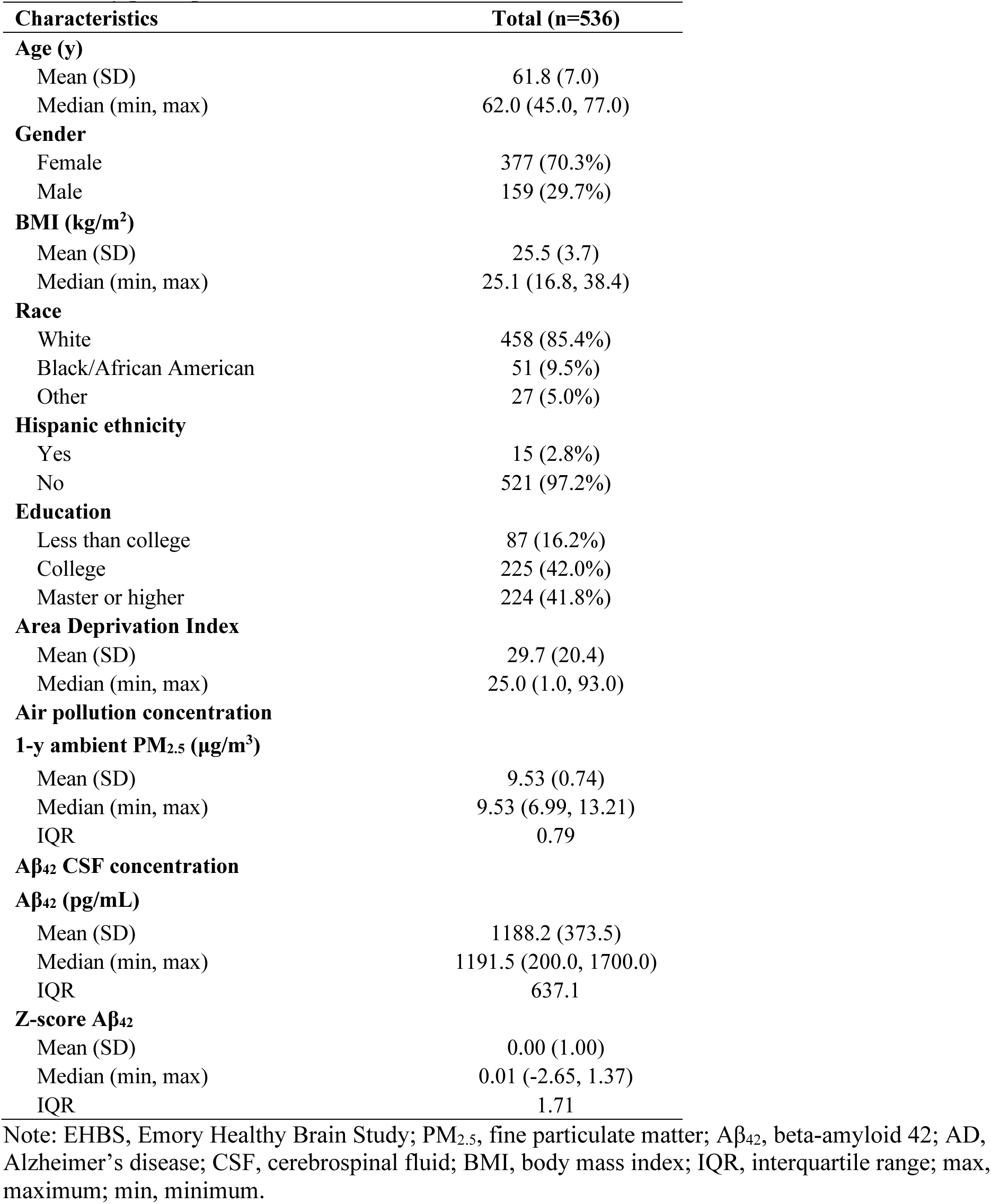
Baseline descriptive characteristics, PM_2.5_ exposure and AD CSF biomarker outcomes for the EHBS study participants.

### 3.2 The total effect of PM_2.5_ on CSF Aβ_42_ concentration

The total effects of PM_2.5_ on CSF Aβ_42_ concentration were estimated using linear regression adjusting for confounders including sex, age, race/ethnicity, educational attainment, BMI, ADI, and three principal components of neighborhood deprivation (**Table S2**). A negative but statistically insignificant association was observed between 1-year average PM_2.5_ exposure and CSF Aβ_42_, with each 1 μg/m^3^ increase in PM_2.5_ corresponding to a -0.074 (95% CI: -0.190, 0.042) decrease in CSF Aβ_42_ z-score. When the analysis was expanded to include a larger sample of EHBS participants not limited to those with DNAm data (N = 1,113), the observed negative association was statistically significant (β = -0.102; 95% CI: -0.179, -0.024), as reported in our previous study[9].

### 3.3 High-dimensional mediation analyses

Standard single-mediator mediation analysis was conducted to assess the CpG site “one-at-a-time” and screen for potential mediators associated with PM_2.5_ exposure and decreased CSF Aβ_42_ levels. Of the 661,869 CpGs tested, there were 1,967 showed p-values < 0.05 for both exposure-mediator and mediator-outcome associations. Among these, 898 CpG sites demonstrated a negative natural indirect effect (NIE), consistent with the negative total effect observed between PM_2.5_ exposure and CSF Aβ_42_ concentrations (**Figure S4**). These 898 CpGs were advanced to downstream high-dimensional mediation analyses using five methods: DACT, HIMA1, HIMA2, HDMA, and MedFix (**Figure 1**). We classified a CpG site as “active” if its estimated mediation effect was non-zero and its raw p-value for significance testing was <0.05 in the high-dimensional mediation analyses. Among the 898 CpG sites analyzed, DACT identified 111 active sites, HIMA1 identified 13, HIMA2 identified 9, HDMA identified 5, and Medfix identified 4, yielding a total of 133 unique CpG sites (**Table S3**). Nine CpG sites were identified as active by two of the methods and were considered noteworthy mediators in our analysis pipeline (**Table 2**). No CpG site was identified as active by more than two methods. The estimated mediation effects for these nine sites were consistent across methods in both effect size and direction, all exhibiting negative effects in line with the expected direction of the total effect (**Table 2**). Based on chromosomal annotation, six CpG sites were located within or near annotated genes: cg0053427[*SLC7A11*], cg11676342 [*BEND7*], cg17904786 [*GTF3A*], cg05532093 [*NFATC1*], cg16891376 [*FLT3LG*], cg05352541 [*PITPNB*]. The remaining three CpG sites were intergenic: cg21682780, cg23129342, and cg10285526.

**Table 2.** Estimated mediation effects and the raw joint significant test p-value of noteworthy CpG sites identified by at least two high-dimensional mediation analysis methods (*DACT*, *HIMA1*, *HIMA2*, *HDMA*, *MedFix*) for the associations between PM_2.5_ exposure and AD CSF biomarker Aβ_42_ concentrations^a^.

| CpG<sup>b</sup> | Chr | Pos | Gene | DACT |  | HIMA1 |  | HIMA2 |  | HDMA |  | MedFix |  |
| --- | --- | --- | --- | --- | --- | --- | --- | --- | --- | --- | --- | --- | --- |
|  |  |  |  | Estimate | p-value | Estimate | p-value | Estimate | p-value | Estimate | p-value | Estimate | p-value |
| cg21682780 | 1 | 200,211,351 | Intergenic | -1.69E-02 | 0.014* | -1.24E-02 | 0.042* | —<sup>c</sup> | — | — | — | — | — |
| cg00534274 | 4 | 139,163,341 | SLC7A11 | -1.86E-02 | 0.071 | -1.97E-02 | 0.007* | -7.30E-03 | 0.222 | -1.36E-02 | 0.027* | — | — |
| cg11676342 | 10 | 13,516,135 | BEND7 | -1.72E-02 | 0.026* | — | — | — | — | — | — | -1.24E-02 | 0.041* |
| cg17904786 | 13 | 27,998,438 | GTF3A | -1.77E-02 | 0.116 | -6.89E-03 | 0.044* | — | — | — | — | -1.03E-02 | 0.044* |
| cg23129342 | 15 | 39,283,510 | Intergenic | -1.61E-02 | 0.474 | -8.54E-03 | 0.045* | — | — | -9.40E-03 | 0.045* | — | — |
| cg10285526 | 16 | 34,787,199 | Intergenic | -1.69E-02 | 0.024* | — | — | -1.17E-02 | 0.035* | — | — | — | — |
| cg05532093 | 18 | 77,186,817 | NFATC1 | -3.32E-02 | 5.09E-08* | -2.27E-03 | 0.193 | — | — | -1.57E-02 | 0.026* | — | — |
| cg16891376 | 19 | 49,976,438 | FLT3LG | -1.64E-02 | 0.034* | -5.50E-03 | 0.043* | — | — | — | — | — | — |
| cg05352541 | 22 | 28,315,563 | PITPNB;<br>LOC284900 | -2.05E-02 | 0.211 | -5.88E-03 | 0.040* | — | — | -1.39E-02 | 0.032* | -9.59E-03 | 0.066 |

<sup>a</sup>Candidate CpG sites included in the high-dimensional mediation analyses were pre-filtered based on their significant associations with both PM<sub>2.5</sub> (exposure) and CSF Aβ<sub>42</sub> concentrations (outcome, standardized as z-scores). Details of the filtering process are described in the Methods section.
<sup>b</sup>CpG sites with non-zero mediation estimates and significant raw joint significant test p-values from at least two high-dimension mediation analysis methods were considered to have noteworthy mediation contributions. All estimates are adjusted for covariates described in the Methods section. Significant p-values (<0.05) are marked with (\*).
<sup>c</sup>HIMA1, HIMA2, HDMA, and MedFix utilized panelized regression to select active CpG mediators. Missingness in the table indicate that the estimated CpG mediation contribution was zero in the respective model.

We further validated these nine CpG sites using causal mediation analysis to evaluate the significance of their NIEs under settings that accommodated exposure-mediator (E-M) interactions and accounted for potential mediator-mediator influence within the causal pathway[43, 66]. When each CpG site was evaluated individually, eight of the nine demonstrated significant negative NIEs (**Figure 3**), while E-M interaction effects were not significant for any CpG tested. The eight CpG sites with significant NIEs included cg21682780 [*intergenic*], cg00534274 [*SLC7A11*], cg11676342 [*BEND7*], cg17904786 [*GTF3A*], cg10285526[*intergenic*], cg05532093[*NFATC1*], cg16891376 [*FLT3LG*], and cg05352541 [*PITPNB*]. All effect estimates reflect changes in the CSF Aβ_42_ z-score per 1 ug/m^3^ increase in PM_2.5_ exposure. The NIE estimates (95% CI) for the eight significant CpG sites, ranged from -0.015 (- 0.036, -0.0008) for cg21682780 [*intergenic*] to -0.029 (-0.062, -0.0069) for cg05532093 [*NFATC1*]. All NIEs were smaller than the natural direct effects (NDEs) but exhibited greater precision. When all nine CpG sites were considered simultaneously as multiple mediators, the estimated joint NIE increased to -0.115 (95% CI: -0.215, - 0.101) (**Figure 3**), while E-M interaction effects were again not significant.

**Figure 3.**
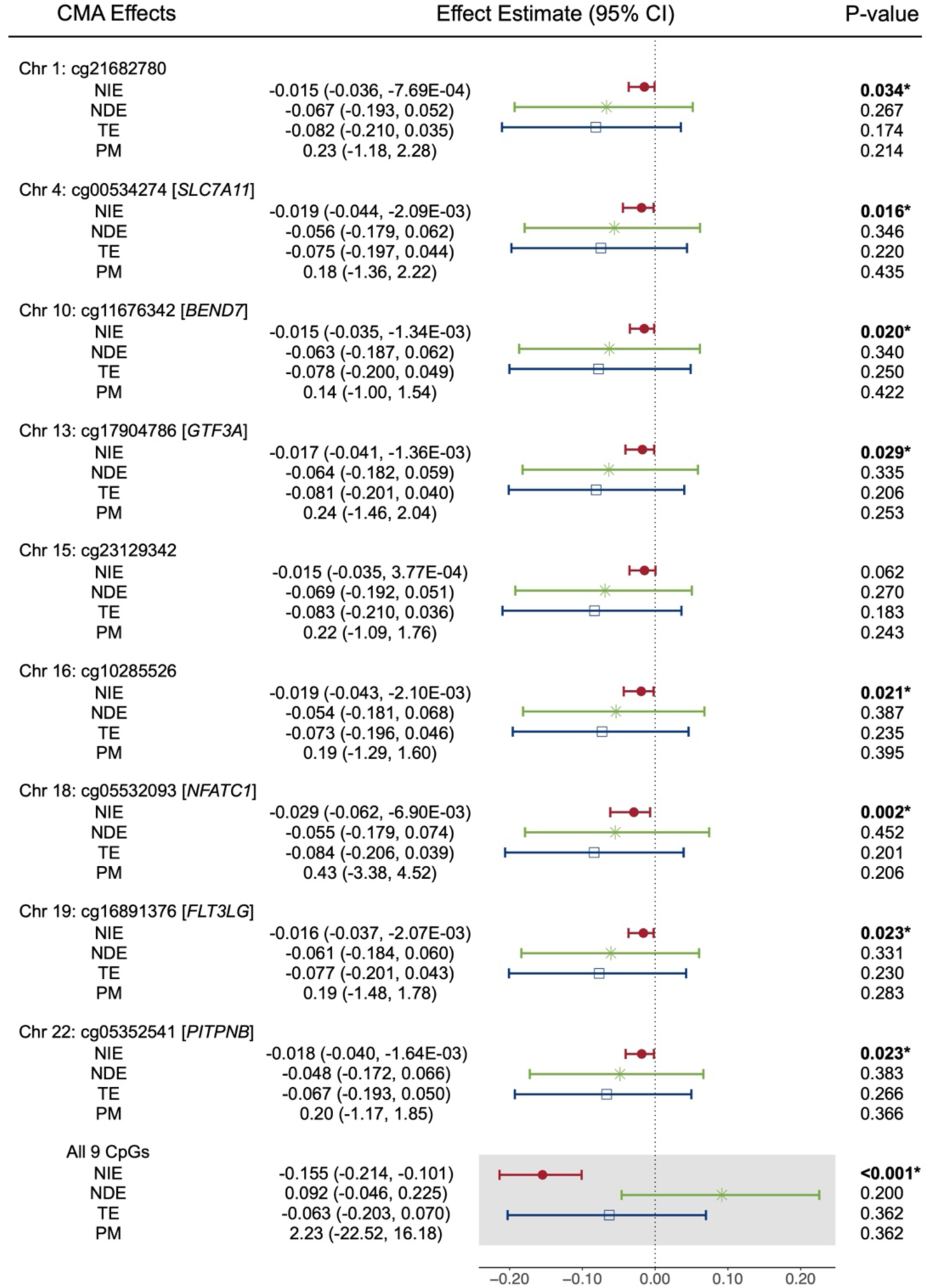
Causal mediation analysis for the association between PM_2.5_ (exposure) and AD CSF biomarker Aβ_42_ concentrations (outcome, standardized as z-scores) using noteworthy CpG sites identified through high-dimensional mediation analysis methods. The figure presents estimates for the natural indirect effect (NIE), natural direct effect (NDE), total effect (TE), and proportion mediated (PM) for selected CpG sites, assessed both individually and jointly. Effect estimates represent the change in Aβ_42_ z-score concentrations per 1ug/m^3^ increase in PM_2.5_ exposure. Significant indirect effects are marked with (*), while no significant direct or total effects were observed.

### 3.4 Sensitivity analyses

To evaluate the robustness of our finding, we repeated the causal mediation effect of the nine noteworthy CpG sites, restricting the study population to White participants (N=458) (**Table S1, Figure S5**). Seven CpG sites retained statistically significant NIEs, including cg21682780 [*intergenic*], cg11676342 [*BEND7*], cg17904786 [*GTF3A*], cg10285526 [*intergenic*], cg05532093 [*NFATC1*], cg16891376 [*FLT3LG*], and cg05352541 [*PITPNB*]. Their NIE estimates (95% CI) ranged from -0.014 (-0.037, -0.0003) for cg21682780 [*intergenic*] to -0.022 (-0.053, -0.0014) for cg05532093 [*NFATC1*] (**Figure S6**). When considering all nine CpG sites jointly, the estimated joint NIE was -0.150 (95% CI: -0.212, -0.095), which was similar to our main analysis.

### 3.5 Look-up of noteworthy CpG sites in cross-tissue databases and pathway enrichment analysis

We performed a blood-brain concordance analysis for the eight CpG sites identified from the high-dimensional mediation analyses that also exhibited significant NIEs in causal mediation analysis, to evaluate how well DNAm patterns in peripheral blood reflect those in brain tissue. Using the BECon tool, four CpG sites demonstrated the highest percentile mean correlations: cg17904786 [*GTF3A*] (90%), cg16891376 [*FLT3LG*] (90%), cg10285526 [intergenic] (75-90%), and cg05532093 [NFATC1] (50-75%) (**Table S4**). The Gene Expression Omnibus Database [Accession code GSE111165] confirmed the significant blood-brain concordance for two of these CpG sites (cg10285526 [intergenic] and cg16891376 [*FLT3LG*]) (**Table S5**).

A gene ontology (GO) analysis was conducted for the 898 CpG sites filtered from the single-mediator analysis (**Figure 1**). After correcting for multiple testing (FDR < 0.05), no GO terms or KEGG pathways exhibited significant overrepresentation of genes containing differentially methylated CpGs. The 295 nominally significant GO terms (raw p<0.05) and the top 20 KEGG pathways were summarized in the Supplement (**Table S6, S7**). The highest-ranking pathway identified was the arginine and proline metabolism pathway, with three differentially methylated genes (*ALDH9A1*, *P4HA3*, *P4HA2*) detected by one high-dimensional mediation methods (**Table S3**).

## 4 Discussion

In this study, we conducted a multi-stage high-dimensional mediation analysis to investigate whether differential DNAm levels in blood mediate the previously identified association between PM_2.5_ exposure and AD CSF biomarker Aβ_42_ in 536 cognitively normal EHBS participants[9]. Using an analytical pipeline incorporating single-mediator analysis and multiple high-dimensional mediation methods, we identified nine noteworthy CpG sites mediating the adverse effect of PM_2.5_ on Aβ_42_. Causal mediation analysis confirmed significant mediation effects for eight CpG sites, and weak correlation among the nine CpGs suggests independent mediation effects (**Figure S7**). Six CpG sites were annotated to genes (*NFATC1*, *SLC7A11*, *FLT3LG*, *PITPNB*, *BEND7*, and *GTF3A*) related to neurodegeneration and air pollution stress responses, suggesting their roles as biologically plausible mediators. To our knowledge, this is one of the first high-dimensional mediation studies examining DNAm as mediators between PM_2.5_ exposure and AD biomarkers in a cognitively normal cohort.

Among the CpGs identified, cg05532093 [*NFATC1*] was the most significant and robust mediator (**Figures 3, S6**). *NFATC1* encodes a transcription factor regulated by the Ca²⁺-dependent protein phosphatase calcineurin (CaN), functioning through the CaN/NFAT signaling pathway[73]. This pathway regulates pro-inflammatory genes that regulate immune response, including cytokines *IL-6*, *TNF-α*, and the apoptotic factor *FasL*[74]. While cg05532093 itself has not been previously linked to AD, several CpGs within *NFATC1* have been associated with AD-related CSF biomarkers, including Aβ_42_, Aβ_38_, p-tau, and neurofilament light chain, in the blood-based EWAS from the European Medical Information Framework for Alzheimer’s Disease (EMIF-AD) study[75]. Brain-based EWASs have also reported differentially methylated sites within *NFATC1* in association with Braak staging [22, 71, 76]. In line with the reported cross-tissue association between *NFATC1* and AD, our blood-brain concordance analysis showed that cg05532093 exhibited moderate correlation (50th–75th percentile) between blood and brain DNAm (**Table S4**), suggesting that its methylation in blood may reflect similar epigenetic changes in the brain. Additionally, multiple *NFATC1* CpGs have been associated with air pollution exposure in blood-based EWAS, including cg06937978 and cg18092363, which were linked to PM_2.5_[32, 33] and NO_2_[77]. Biochemical and molecular studies also suggest CaN/NFAT signaling can be a mediator of Aβ-induced neurotoxicity in both early and advanced stages of AD progression[73, 78], and inhibitors targeting this pathway (e.g., cyclosporine A, tacrolimus) has shown promise in alleviating AD symptoms [74].

Another significant mediator, cg00534274, is located in the first exon of *SLC7A11*, encoding a subunit of the cystine–glutamate antiporter system Xc-, which is critical for antioxidant defenses. DNAm at this site has been linked to systemic inflammation via C-reactive protein (CRP) levels in EWASs[79], suggesting a link between PM_2.5_-induced oxidative stress and epigenetic regulation of *SLC7A11*. This antiporter also modulates glutamate release, an essential process in excitatory neurotransmission that is disrupted in neurodegenerative diseases[80]. Other CpGs within *SLC7A11*, including cg07661704 and cg06690548, have been associated with AD and Parkinson’s disease in blood-based EWASs[75, 81]. A recent study reported increased *SLC7A11* mRNA and pLG72 (D-amino acids oxidase modulator) proteins levels in plasma from AD patients, with the combination of these biomarkers showing high specificity and sensitivity for AD detection (AUC=0.882)[82].

Cg11676342 [*BEND7*] was another noteworthy mediator. *BEND7* encodes a protein involved in protein-DNA and protein-protein interactions. One CpG (cg07995537) within *BEND7* was previously associated with abnormal CSF t-tau levels in a blood-based EWAS in differentiating AD, MCI patients to healthy controls[75]. Bioinformatics study have identified *BEND7* as part of an AD-specific molecular network[83], though its role in AD remains poorly understood. Additionally, *BEND7* CpGs have been associated with BMI[84, 85], CRP levels[79], rheumatoid arthritis[86], but its link to PM_2.5_ remains unexplored.

Cg17904786 [*GTF3A*], located in the promoter region of a gene encoding General Transcription Factor IIIA, was also a significant mediator. This CpG site has been associated with age[87] and rheumatoid arthritis[86], suggesting its relevance to age-related processes. In the context of AD, another CpG in GTF3A (cg22180069) was among the 1000 differentially methylated sites associated with CSF Nfl levels in AD patients[75]. Transcriptomic studies have also identified *GTF3A* as differentially expressed in AD brains with varying severities, suggesting a role in AD progression[88].

Cg16891376 [*FLT3LG*], located in the promoter region of a gene encoding a cytokine involved in dendritic cell development, was also a noteworthy mediator. A recent study identified *FLT3LG* as one of the top-ranked CSF immune activation markers associated with neuronal injury in preclinical AD[89]. These findings suggest that changes in CSF FLT3LG and its DNAm status may occur during the presymptomatic stages of AD pathogenesis[89, 90].

Cg05352541 [*PITPNB*], encoding a phosphatidylinositol transfer protein, plays critical roles in lipid signaling and membrane dynamics. Two CpG sites within *PITPNB* has been associated with tobacco smoking in blood-based EWASs[91, 92]. While its connection to AD remains underexplored, transcriptomic analyses have identified *PITPNB* among the top 300 hippocampal genes potentially involved in AD neuropathology[93]. Additionally, a proteomic study using brain tissue-derived dataset reported *PITPNB* as a predictive biomarker distinguishing asymptomatic AD from controls[94].

Our KEGG pathway enrichment analysis identified the arginine and proline metabolism pathway as the top-ranked pathway associated with PM_2.5_ exposure and CSF Aβ_42_ levels. This pathway have been implicated in AD progression, with metabolomics studies detecting alterations in arginine and proline metabolism across multiple tissues, including the brain, CSF, and plasma[95, 96]. Notably, this pathway is also a well-established sensitive marker of air pollution and traffic-related PM exposure[97, 98], suggesting <u>perturbation</u> in arginine and proline metabolism as a potential link between PM_2.5_-driven oxidative stress and AD-related neurodegeneration.

This study has several notable strengths. First, we employed a rigorous analytical pipeline that integrated single-mediator analysis, multiple high-dimensional mediation methods, and causal mediation analysis. Previous studies using multiple high-dimensional methods have reported low concordance across methods and limited statistical power to detect mediation effects[34, 54, 55]. To address these challenges, we followed recommendations from Clark-Boucher et al. (2023) to compare CpG sites results across five distinct methods (DACT, HIMA1, HIM2, HDMA, and MedFix) with nominal significance (raw p-value <0.05)[44]. We considered CpG sites as noteworthy active mediators if they were identified by at least two methods, which reduced the risk for false discoveries. To further validate our findings, we assessed the indirect effects of these CpG sites using causal mediation analysis that accounted exposure-mediator interactions[42, 43], confirming statistically significant mediation effects for eight of them. This approach added robustness and confidence to our results. We also estimated the joint mediation effects of all nine identified CpG sites, providing an overall view of their total mediation effect. Second, the EHBS is among the largest prospective cohort studies that include CSF samples from cognitively normal individuals[45]. The comprehensive nature of our outcome assessment—a highly invasive and difficult-to-obtain biological fluid— offers a unique and valuable opportunity to explore potential associations between PM_2.5_ exposure, differential DNAm and AD CSF biomarkers in cognitively normal individuals. Third, our use of a high-resolution PM_2.5_ exposure model enabled precise characterization of spatial variability in individual exposure, reducing potential measurement error.

However, our study has several limitations. First, our analyses were constrained by a relatively small sample size (N=536), which may have limited the statistical power to detect DNAm mediation effects on CSF Aβ_42_ levels in cognitively normal EHBS participants. The limited sample size might have also reduced our ability to detect total effects between ambient PM_2.5_ exposure and CSF Aβ_42_ concentrations. Notably, while a negative but non-significant total effect was observed in the current analysis, a significant negative total effect was detected when the analysis was expanded to the larger EHBS participant sample not restricted to availability of DNAm data (N=1,113)[9]. Future studies with larger sample size are essential to confirm and validate these findings. Second, DNAm signatures are tissue- and cell-type specific, and DNAm in brain tissue would be more directly relevant for studying CSF AD biomarkers. Although CSF surrounds the brain, it has been reported ∼80% of CSF proteins originate from blood plasma that crosses the blood-brain barrier[99]. Thus, it is plausible that the DNAm alterations we observed in blood could mediate CSF transcriptomic and proteomic changes. Importantly, peripheral tissue samples such as blood are far more feasible to collect than brain tissue, enabling the possibility of gathering longitudinal samples to investigate temporal changes in DNAm, AD biomarkers, and cognitive decline. Third, the temporal sequence between mediators (DNAm changes) and outcome (CSF Aβ_42_ concentrations) could not be clearly defined, as both biospecimen were collected at the baseline visit. However, as EHBS is an ongoing cohort, future data may allow for a better longitudinal study design to address this limitation. Finally, while our findings suggest a potential mediation role of neuroinflammatory pathways in the association PM_2.5_ exposure and CSF AD biomarkers, the study did not directly measure the proinflammatory factors or the expression of related genes in blood or CSF. Future studies incorporating multi-omics approaches —including transcriptomics, proteomics, and metabolomics— are warranted to validate and expand upon our findings.

## 5 Conclusion

Using a combination of single-mediator analysis, high-dimensional mediation analysis, and causal mediation analysis, this study identified differential DNAm in blood as a significant mediator of the association between PM_2.5_ exposure and CSF Aβ_42_ concentrations in the EHBS cohort of 536 cognitively normal individuals. Nine differentially methylated CpG sites were identified as noteworthy mediators, with their corresponding genes implicated in pathways related to neuroinflammation and neuroinflammation-mediated necroptosis. These findings provide valuable insights into the biological mechanisms underlying PM_2.5_ toxicity during the early stages of AD pathogenesis. They also highlight the potential utility of blood DNAm changes as accessible biomarkers for detecting AD-related alterations and environmental exposures. While the relatively small sample size and the current lack of a universally accepted gold-standard approach for high-dimensional mediation analysis suggest this study serves as an initial investigation, the results are novel and promising. Further studies are warranted to replicate and expand these results, enabling a deeper understanding of how DNAm and other biological pathways mediate the impact of air pollution exposure on AD development.

## Data Availability

All data produced in the present study are available upon reasonable request to the authors

## Acknowledgments

We gratefully acknowledge the research volunteers and staff of the Goizueta Alzheimer’s Disease Research Center at Emory University, and Emory Healthy Brain Study for their participation and contributions. This work was supported by I01 BX003853 (APW), I01 BX005686 (APW), IK4 BX005219 (APW), P30 AG066511 (AIL, JJL), R01 AG056533 (APW, TSW), R01 AG070937 (JJL), R01 AG072120 (APW, TSW), R01 AG075827 (APW, TSW), R01 AG079170 (AH, TSW), U01 AG046161 (AIL), U01 AG061356 (AIL), U01 AG061357 (AIL), U01 AG088425 (AH, TSW, DL), R01AG087250 (AH, DL)

## Consent Statement

The Emory Healthy Brain Study was approved by the Institutional Review Board of Emory University Medical Center. All participants provided written informed consent.

